# Aging, Care and Dependency in Multimorbidity: How Relationships Affect Older Bangladeshi Women’s Use of Homecare and Health Services

**DOI:** 10.1101/2020.06.19.20126078

**Authors:** Mohammad Hamiduzzaman, Stacy Torres, Amber Fletcher, Md. Rezaul Islam, Jennene Greenhill

## Abstract

Relationships are multidimensional, and we know little about how different facets of relationships affect how older patients’ with multimorbidity use homecare and health services. Social gerontology literature emphasizes the importance of care settings, gender inequalities, availability of health services, and affordability. However, the diversity of relationships and associated dependency in elder care remains underassessed. This qualitative study combining a demographic survey with interviews explores the relationship experiences of older women (age 60 years and over) with multimorbidity in homecare and health services utilization. Researchers contacted the Civil Surgeon of Sylhet District in Bangladesh to recruit study participants and conducted 33 interviews [11 staff members and 22 older women with multimorbidity]. Three domains of Axel Honneth’s theory of recognition and misrecognition [i.e. intimate, community, and legal relationships] underpin study findings. Data were analysed using critical thematic discourse analysis. Four themes, including seven relationship dimensions, emerged: the nature of caregiving; intimate affairs [marital marginalization and parent-children-in law dynamics]; alienation in peer-relationships and neighbourhood [siblings’ overlooking of women’s rights and needs, neighbourhood challenges such as ageism, and gender inequality in interactions]; and legal connections [ignorance of rights and missed communication]. Marginalization in family relationships, together with poor peer supports and a misrecognition of care needs from service providers, resulted in a lack of quality care for older women with multimorbidity. Understanding the complexities of older women’s relationships may assist in policy making with better attention to their health needs and deepen understanding of how gender inequality intersects with the cultural devaluation of older adults to reduce their well-being. Staff training on relationship building and counselling services for family caregivers and kin are essential to improve the quality of care for these women.

## Introduction

Relationships are integral to the quality of older multimorbid patients’ homecare and health services usage [1, 2]. Many older Bangladeshi women have multimorbid, chronic health conditions that affect physical capacity and daily living activities, such that they require ongoing homecare and medical care support [3-5]. Multimorbid patients’ care involves: (i) regular medical visits and treatments provided by clinicians and (ii) non-medical care including in-home assistance, financial support, accompaniment to clinical assessments, management of treatments and rehabilitation, generally supported by family members, relatives and neighbours [6]. Social gerontology studies suggest that a care-focused relationship approach by family, peers, and clinicians, based on recognition, compassion, active listening, and sensitivity to older patients’ needs and preferences, can improve the health and well-being of older women with multimorbidity [7, 8]. In relationships undergoing deliberate depreciation, older women often face threats to their emotional attachments and also grapple with health consequences of ignorance and isolation, which include implicit exclusion from family and health systems and ageism in community and medical settings [9].

In developing countries, older women’s intimate relationships tend to be circumscribed and their socioeconomic and cultural relationships are transitional in nature. Studies that have examined separately their familial relationships, neighbourhood and clinical communication experiences, highlight three realities: (i) dependency on family members for homecare and financial assistance; (ii) neighbouring relationships for care and advice; and (iii) the significance of clinicians to encourage service use [10, 11]. We know little about how the relationship experiences of older women multimorbid patients shape their search for ongoing medical care and in-home care management. It is assumed that they require increased care and first seek assistance from family members, peers, and neighbours [12]. However, prior research finds when family members perceive their condition to worsen and require financial support, isolation from family and a range of social relationships occurs [12, 13]. Detachment also occurs for older women in friendship and neighbourhood relations due to limited mobility and male dominance in family, social, and clinical interactions [14].

To understand the quality of relationships for older women with multimorbidity and how these relationships shape care support in a developing country like Bangladesh, we draw on a focused literature review, theoretical explanations, and observation of relationship pathways and mechanisms that influence health and well-being [1,6]. Attachment in diverse relationships, including family care [e.g. love and care], social support [e.g. advice and friendship] and clinical interactions [e.g. recognition and positive approach], can influence psychosocial, behavioural, and physiological well-being [11, 14]. Relationship variations emerge due to excessive demands, progression of care burden, geriatric knowledge of carers, caregivers’ empathy for caregivers, and available social resources [15, 16]. Need for growing financial support from family members also contributes to poor psychosocial outcomes, especially in terms of healthcare access and the increased time in care [17, 18].

We begin with an overview of older Bangladeshi women’s living circumstances, health, and well-being. We identify affection, reliance, care, and equality as four key features that influence older women’s relationships with family, community, and clinicians. Study findings generate knowledge about participants’ experiences and views on relationships and associated dependency that affects their homecare and health services use. We conclude with a discussion of how different relationships facilitate women’s health and well-being or promotes inequality and discomfort. This study contributes to scholarship on aging, care, and dependency by expanding knowledge of complexities in elder relationships and diverse relationship structures that help older women connect and secure informal care yet may also hinder them from obtaining care as gender and age inequalities intersect in late life.

## Older women and quality of life in Bangladesh

The older adult population (i.e., aged 60 and over) is increasing in Bangladesh. Increased longevity coupled with high rates of multimorbidity and chronic illness cause specific care needs that family caregivers and healthcare systems have yet to adequately address [1]. Approximately 70% of older women live in rural areas and are more likely than their urban counterparts to depend on families to manage medical treatments [6]. Only ten in every 10,000 women will seek healthcare at local public and private hospitals, and their hospitalization and emergency presentation rate remains as low at 5% of all in-patients admissions [8, 15]. Limited healthcare use by older women is shaped by social relationships and interacting socioeconomic, cultural, and political structures. Existing literature indicated the significant role of family and the health system in shaping homecare and healthcare use, as older women tend to downplay their illnesses, delay treatment, and depend on lay or traditional healers who may exploit them and/or provide inappropriate care [1, 6, 8, 16]. Other interconnected barriers for older women’s healthcare include a lack of services in rural areas, low levels of education and health literacy, and gendered economic inequality — for example, Muslim women inherit only 1/8 of a deceased husband’s property, and married Hindu women cannot inherit their parents’ property, promoting life-long dependency due to the extreme lack of public income supports for this group [15, 17]. This qualitative study enriches scholarly understanding of the complex interplay of familial, social, and clinical relationships that shape homecare and healthcare service use for older women with multimorbidity in Bangladesh.

## Materials and methods

### Research design

This qualitative study draws on a focused understanding of theoretical and practical explanations of older women multimorbid patients’ care needs, a demographic survey, and semi-structured, in-depth interviews.

### Ethics

The Social and Behavioural Research Ethics Committee at Flinders University (*Project No. 6705*) granted ethics approval for this study. Formal data collection permission was obtained from the Directorate General of Health Services, Department of Primary Healthcare and Sylhet Civil Surgeon Office of Ministry of Health and Family Welfare of Bangladesh prior to submitting the ethics application, prepared according to the guidelines of the National Health and Medical Research Council, Australia.

### Theoretical framework

We used the principles and domains of Axel Honneth’s (1996) theory of recognition and misrecognition to provide theoretical underpinnings for study findings [18, 19]. This theory emerged from a relationship-focused research philosophy and facilitates the identification of relationship complexities that influence the use of homecare and health services. The framework identifies three domains in human relationships: intimate relationships, a legal framework, and the community environment. Each domain provides direction about how recognition or misrecognition occurs in relationships [Table 1]. We applied them in analysis and discussion sections to explain the relationship structures of older women with multimorbidity and how these relationships impact their homecare and health services usage.

**Table 1.** Relationship domains and forms of recognition

| Domains | Recognition and misrecognition areas | Forms of recognition |
| --- | --- | --- |
| Intimate | Intimate relationship [e.g. spouse] | Love and positive attitudes to each other |
|  | Family relationship [e.g. children, grandchildren] | Mutual recognition and respect of needs and preferences |
| Legal | Laws and rules [e.g. constitutional, health policy] | Legal validation of human rights |
|  | Institutional relationships [e.g. health staff - client] | Role play as a social actor |
| Community | Social structures [e.g. organizations] | Agreed body of rights |
|  | Peer relationships [e.g. neighborhood] | Mutually share the values and one's personalities and abilities are appreciated and supported by others |

### Study setting and participants

We approached health staff in community clinics/hospitals and community-dwelling older women in the North-Eastern region [i.e. three residential communities of Sylhet District: Daldali, Hiluasura, and Gouri Pur] of Bangladesh, by contacting the district Civil Surgeon. Information sheets and consent forms with reply-paid envelopes were provided to the Civil Surgeon for distribution among staff in formal meetings and to women in yard-gatherings. Inclusion criteria for staff recruitment encompassed the following: working in Sylhet Sadar Upazila; experience in providing care to older women with multimorbidity; and willingness to participate in an interview. Older women recruited possessed the following inclusion criteria: lived in the selected residential communities; self-reported multimorbid patients; able to provide consent; and interested in participating in a demographic survey and interview. We informed participants that they could withdraw at any time and would experience no adverse immediate or future effects upon their care provision/access, and that we would use pseudonyms in reporting.

### Data collection

Eleven staff and 22 older women met inclusion criteria. Staff [clinicians, pharmacists, and public health assistants] participated in face-to-face, audio recorded, in-depth interviews, and older women with multimorbidity took part in a short demographic survey and interview with a similar format. Thirty-five older women expressed interest in participating, but 13 women were excluded as not being multimorbid patients. Interviews began with demographic questions about age, education, income, marital status, household size, self-reported health conditions, and service usage. Open-ended questions followed by prompts guided discussion about family members’ understanding and approach to women’s care needs, shared values and relationships with peers and neighbors, and health staff’s approach and support. Interviews were conducted in Bangla (i.e. native language), and the first author transcribed and translated interview audio-recordings into English for analysis.

### Analysis of data

We used a critical thematic discourse technique in data coding and thematization. As thematic analysis focuses on the identification of patterned meanings of surface reality [20, 21], this technique was integrated into the data analysis process to provide an understanding of discourses and dialectical relationships [22]. NVivo 11 software facilitated analysis. Analysis began with gaining familiarity with data by reading and re-reading transcripts and applying automated coding. At initial coding, all potentially applicable determinants, actions, events, and meanings were labelled as open codes. Focused coding followed, conducted according to the domains of Honneth’s (1996) theory of recognition and misrecognition, and clustered codes into nodes. The first author reviewed the nodes/clusters in relation to audio recordings and transcripts for searching candidate sub-themes and themes. The research team cross-checked codes, nodes, and candidate sub-themes to name and define sub-themes and themes. To retain reliability and validity, themes that emerged from the researcher’s coding and categories were discussed and reviewed in the project’s fortnight meetings. Group discussion also considered defining, naming, and contextualising themes to represent practical realities about how relationships affect care for women multimorbid patients.

### Data reporting

To report data, the Consolidated Criteria for Reporting Qualitative Research checklist was employed [23]. As such, sub-themes and themes were presented with the participants’ quotations to exemplify the findings.

## Results

### Baseline demographics

The demographic characteristics of the women appear in Table 2. The mean age of women was 72 years (ranged from 60 to 100 years). Of the 22 women examined in three villages, 68% had no formal education, and 16 women (73%) had no income at all. While 18 women (88%) were widowed, the average household size was four persons and most (15 women) lived in an extended family, such as with their married sons. Women mostly reported physical diseases where the commonly described symptoms included physical weakness, breathing problems, cough, fever, headache, low vision, head infestations, and stomach problems. Most symptoms could be interpreted using medical terminology, such as heart disease, high blood pressure, physical pain (e.g. chest pain, leg pain, back pain, lower back pain), arthritis, diabetes, chronic fever, and gastric issues. 91% (20) of participants experienced at least three diseases and 64% (14) reported four diseases. However, number of visits to local clinics/hospitals was low, including the community clinic (36%), Union Health Centre (23%), Upazila Health Complex (0%), major public and private hospitals (50%).

**Table 2.**
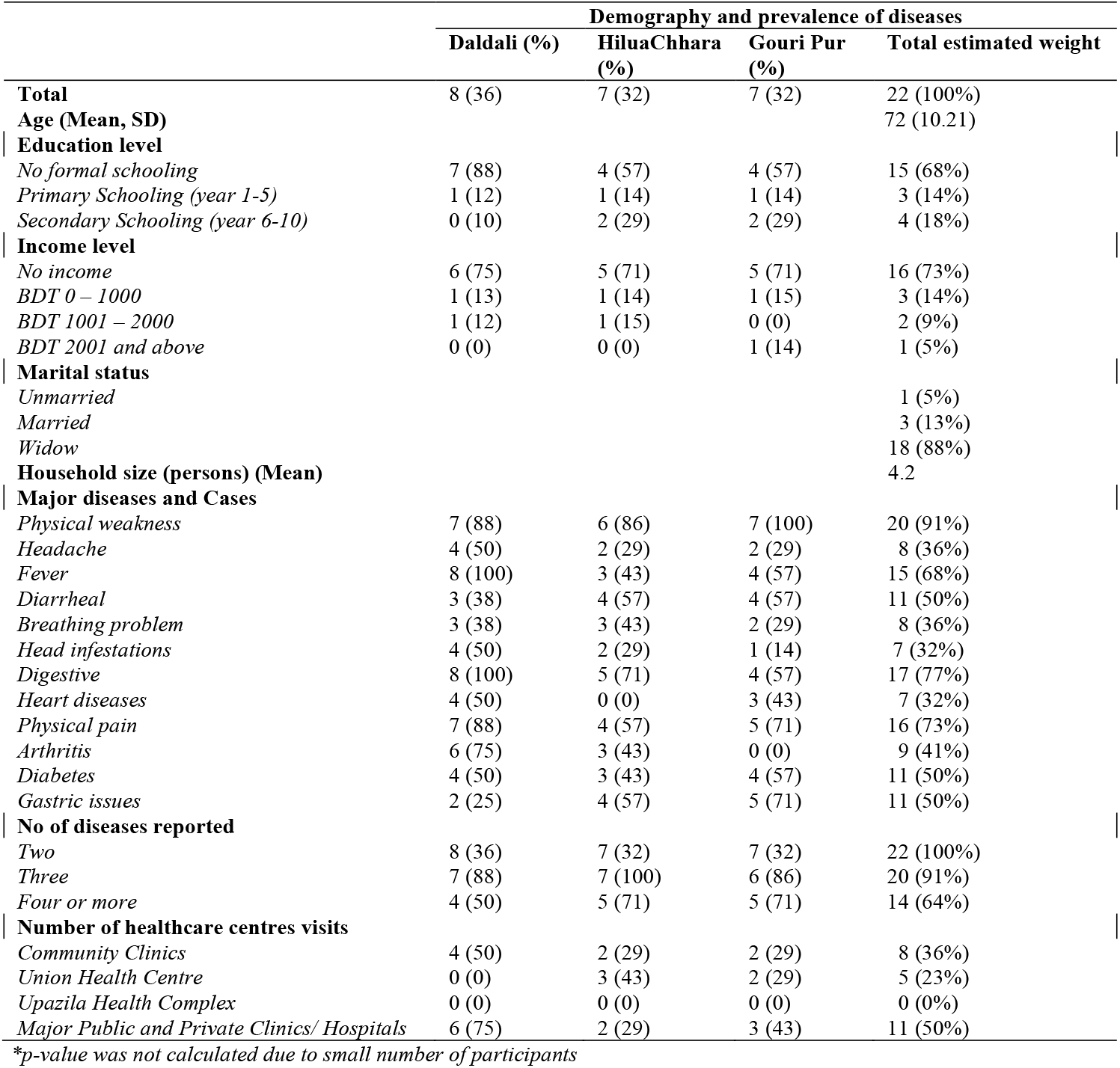
Demographic characteristics and prevalence of diseases among women participants

### Relationship dimensions in multimorbidity

Four main themes, including seven relationship dimensions, emerged from the data: nature of caregiving; intimate affairs [marital marginalization and parent-children-in law dynamics]; alienation in peer-relationships and neighbourhood [siblings’ overlooking of women’s rights and needs, neighbourhood challenges, and gender inequality in interactions]; and legal connections [ignorance of rights and missed communication] [Figure 1].

**Fig 1.**
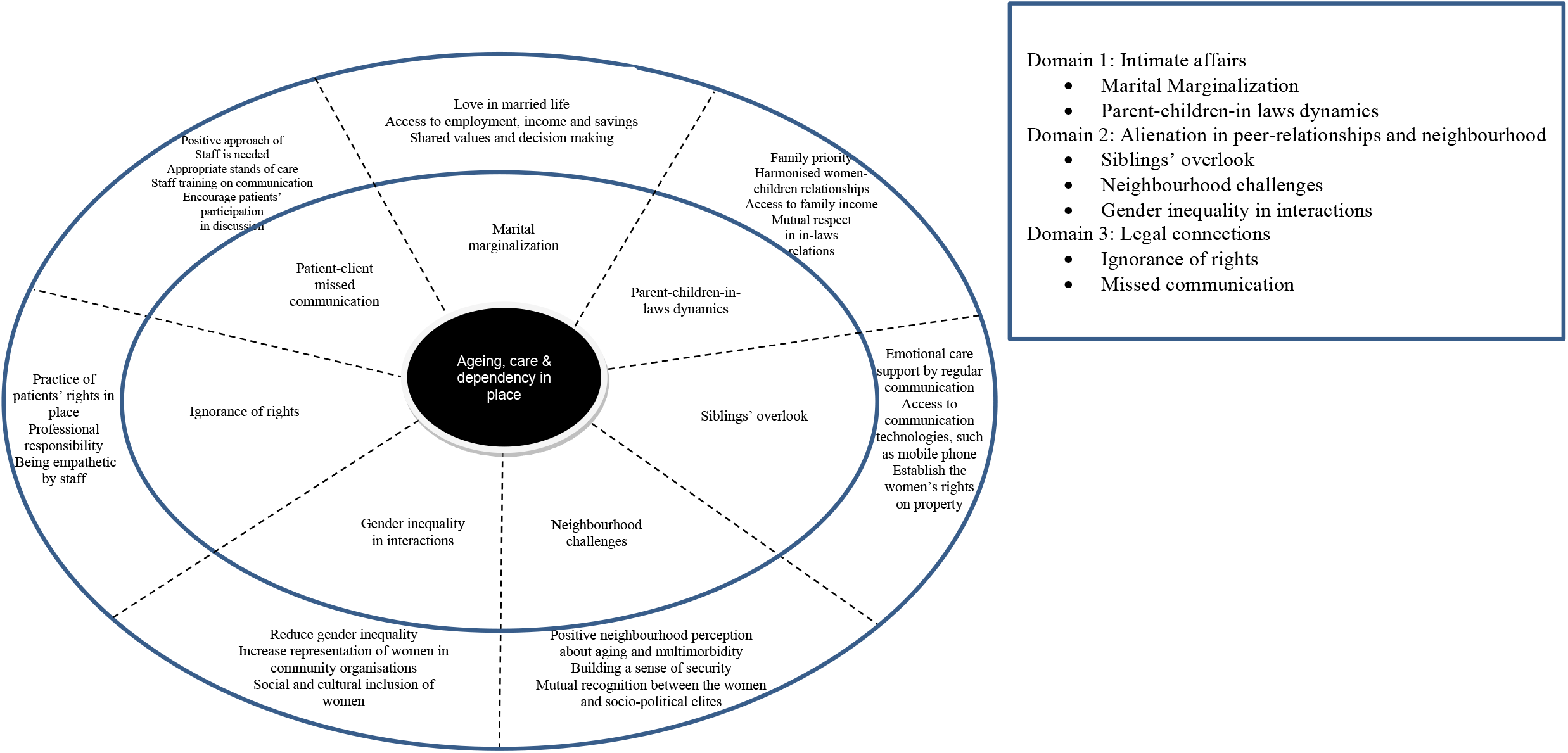
Relationships dimensions into care for older multimorbid women patients

#### Nature of caregiving

The first major theme of ‘nature of caregiving’ demonstrated participants’ homecare and health service needs and caregivers’ involvement in providing care.

Staff described the health and well-being of older women as consisting of physical and psychological symptoms and diseases, and chronic with multiple morbidities. Similarly, twenty older women reported at least three diseases, indicating the widespread prevalence of multimorbidity and chronic conditions that required intensive home and hospital care.

> *According to me, most elderly women are suffering from at least two diseases out of hypertension, diabetes, arthritis, and gynaecological disorders. You will find them physically weak because of malnutrition. They also experience mental disorders like menopausal syndrome, depression, which largely affect them. Moreover, isolation is very common in this area as young family members do not spend time with them*. [Clinician 11; p.110:19-22]
>
> *I have problem with my eyes and ears. I cannot see and cannot hear properly. You have to be loud in asking questions. I have pain in my legs, so I cannot walk or stand alone for a long time. Sit down all the time at one place – how hard it is? Please do something for me, are you a doctor? I am suffering a lot because of my joints’ pain* [arthritis] *and lower back pain*. [Older Woman 2; p.131:18-21]

Participants described care as mostly in-home assistance and managing treatments outside home. They stated that daughters and daughters-in-law provided care related to food intake, rest, timely use of medication, massage during physical pain, emotional care, organise bed for sleep, bathing, and sharing care needs.

> *I stay at home and my daughters-in-law look after me when I become sick. I have two daughters-in-law, who are very good and well-behaved. … Wife of my older son provide food for three-times a day and help me in taking medications at night. When she was pregnant last year, it became difficult for me to have medications on right time, because only she knew that what and when medications should take. You know I feel shy to tell my son about my diseases, but my daughters-in-law are very good to inform my sons about my diseases or if I need anything*. [Older Woman 22; p.315:12-15]

Participants described medical care management as help with transportation, accompanying them to clinics or hospitals, clinical discussions, managing pathological tests and medications, and finances, generally managed by family members, peers, and neighbours.

> *Elderly women in this union are poor and they depend on traditional healing and self-care at home. Women are generally supported by their family members. These elderly women used to be accompanied by male family members when they travel to hospitals or managing cost of visits, tests and or medications. These family members also play an important role in discussion with us*. [Public Health Assistant 7; p.77:13-16]

Both participant groups confirmed family members’ role in meeting women’s needs for homecare and health services.

#### Intimate affairs

The second main theme of ‘intimate affairs’ referred to the dependency of older women with multimorbidity on family. Participants spoke of family relationships in terms of marital relations and parent-children dynamics, and these relationships served as essential sources of homecare and also cast a shadow over health service use.

#### Marital marginalization

Participants posited marital relationships as vital in meeting care needs that arise in late life and affects care support for her. Marriage, particularly feeling unhappy in married life, was associated with financial dependency on husbands and gender inequalities in healthcare decisions.

According to participants, older women with multimorbidity experienced financial dependency on their husbands due to having no to little income because of engagement in unpaid domestic labor rather than formal employment.

> *They used to get money from their husbands. If they have husbands, husbands generally take all financial responsibilities for their wife in families, men earn and spend money. It is a cultural issue and women are only engaged in household activities where they do not get any payment*. [Public Health Assistant 3; p.33:18-21]
>
> *I did not do any job in my life. I only do household works for my family. As Allah gives me the opportunity to take care of my husband and children, I devoted my time and efforts for my family members, and this is the best job I have done. My husband used to spend all money to our family’s monthly expenditure. We cannot save a single penny that makes it hard to visit hospitals when I am in need. I feel shy to request him* [Husband] *to bring medications as I know his capacity*. [Older Woman 21; p.126: 4-6]

Gender differences in household income contributed to women’s dependency, which demoted health and well-being through reduced access to financial and health resources.

All participants described marital decision-making processes as male-dominated; thus, seeking care for a woman with multimorbidity became her husband’s decision, and she had to inform him about her diseases and healthcare needs.

> *My husband used to take all decisions in our family. What are you talking about? Who will take the decision in my family? My elder son lives in capital city ‘Dhaka’ and my younger son live in another village. So, all the decisions are made by my husband*. [Older Woman 1; p.126:8-10]
>
> *They feel afraid about the health problems, because they think about how their husband will react to their health problems*. [Clinician 5; p.51:16-17]

Staff’s description of husbands’ attitudes revealed power relations in marital relationships that negatively affected women’s access to medical care. Such dependency continued even after the husband’s death.

> *Male members of family take decisions for their healthcare, such as husband and then son* [if husband died]. *If they do not have husband, it becomes hard for them to access health services. These women cannot take any decisions by themselves*. [Public Health Assistant 3; p.33:18-21]

#### Parent-children-in law dynamics

Older women with multimorbidity experienced low access to home-based care and health services due to prioritization of young and male family members, breakdown in parent-children relationships, a lack of household financial support in widowhood, and in-law women ties.

Most participants indicated that older women had an obligation to meet the needs of relatives such as sons and grandsons, first, which constrained their ability to access home and clinical care. Consequently, they had to forgo homecare and regular hospital visits, and their own needs ranked as a low priority in the family.

> *When my one son was sick, I had to sell this land for his healthcare. I got 7,000 BDT by selling a land property and spent all money for my son’s treatments. I lost my money and my son both that made me poor. … I did not go for hospitals for my treatments as I had no savings. I also went for XXX medical and XXX medical for my grandson when he was seriously injured*. [Older Woman 7; p. 176:8-11]
>
> *How can I go there for me? My life comes to an end. I have no problem if I do not take medications. I wish that my granddaughters will live healthy. I have sympathy for them. I always ask my daughters-in-law to take care of my granddaughters as they will live in the world*. [Older Woman 8; p.186:23-26].
>
> *Actually, they do not say about their health problems. … It may be because they do not want to share the health problems. Family is first for these women and they always sacrifice for others* [young family members]. [Clinician 9; p.88:7-9]

Analysis revealed that most women experienced a lack of support because of diminishing relationships with sons, daughters, and grandchildren. According to the women and staff, this lack of care grew within the family due to neglect of elder parents by young family members and heavy workloads of relatives that restricted women from timely access to food and healthcare. This breakdown in family relationships resulted in fewer household members who could provide home care and also financial support to women seeking healthcare.

> *My sons are not living with us, and they do not help us. This is a big problem. If my children were good, my family condition could be good. … if they would live with us, they might understand my conditions and take me to the hospitals*. [Older Woman 1; p.124:12-14]
>
> *When we visited door to door, we found elderly women sitting on the floor, and their son and his wife went for work. For example, I saw this today as well. One elderly woman waits for her son and his wife to get food. These women did not get food on the right time because their family caregivers have other priorities* [Public Health Assistant 2, p.19:13-16]

Most participants confirmed the financial dependence of older women on young male family members, especially in widowhood. They described financial dependence on son/s and grandson/s in terms of paying for transportation, clinical visits, pathological tests, and buying medications.

> *In family, all members cannot contribute* [financial support] *for the well-being of the elderly members and they cannot manage time for them to look after. … They keep their mother at home and manage treatments for them at home*. [Clinician 10; p.102:15-17]
>
> *When my sons can manage money, I go for seeing local doctors. If they cannot, I keep patience. What else I can do?* [Older Woman 17; p.260:14-15]
>
> *My sons may give money if they have extra. As they have no extra, they do not give me anything. They have their own families*. [Older Woman 6; p.167:3-4]

Dependence prevented them from making independent healthcare decisions even after their husband’s departure.

Some participants indicated daughters-in-law played a major role in meeting daily care needs. However, they acknowledged a misrecognition of women’s needs and preferences by their daughters-in-law due to relationship conflict.

> *I thought that I could live with my three sons in a joint family. However, it did not happen because my elder son lost lots of money and he has no contribution in the family. He became separated for managing his own family. … If I become sick, my second daughter-in-law used to look after me. I had a good relationship with my daughter-in-law. She used to bring medications and take me to the doctor. She sends food for me when she prepares a good dish*. [Older Woman 5; p.159:15-22]
>
> *There are always issues in the relationships between elderly women and their daughters-in-law. I do not go into the problems, but it is for sure some daughters-in-law want to live as nuclear family and do not want to take responsibility of their mothers-in-law*. [Public Health Assistant 7; p.77:13-16).

### Alienation in peer-relationships and neighbourhood

Similar to family relationships, participants characterised other social contacts as positive and negative. The third major theme of ‘alienation in peer-relationships and neighbourhood’ presented a context whereby older women with multimorbidity faced repression in social relationships due to siblings’ overlooking of women’s rights and needs, neighbourhood challenges, and patriarchal practices in interactions.

#### Siblings’ overlooking of women’s rights and needs

Both participant groups described siblings’ support, such as emotional care and financial support.

Participants described the role of siblings as substantial, especially in providing emotional support to address multimorbid conditions. When the women were asked about their present relations with siblings, they began by discussing how they shared their emotions, thoughts, intentions, and beliefs with sisters in childhood. According to several women, they lost connection with sisters because of their marital relationships but maintained some communication with brothers who occasionally visited them. Breakdown in sibling relations after marriage resulted in a lack of emotional support when women faced broken and isolated family relationships or in-person assistance when they needed to visit local clinics.

> *Immediately after marriage, I thought it could not be possible for me to stay at a different place without my sisters. However, I had to adjust here* [in law’s house] *with my husband and his parents and siblings. It seemed like I lost my sisters and brothers. My sisters moved to other districts because of their marriage and busy with their own life. My brothers still visit me occasionally. As I am sick, they try to communicate and advise me how to get rid of pain. But you know relationships have been changed - no difference between brothers and neighbours. They are busy at work and life – had no time to visit me when I was in hospital for a week. They even did not call me – did not ask me how I went to the hospital. I stay at home alone* [after husband’ death] *when my sons and daughters in law go for work and want to talk with someone. As I do not have mobile phone, I cannot talk with anyone. If I feel pain, I cannot share*. [Older Woman 22; p.289:5-14]

Participants also identified another aspect of siblings’ relations related to property rights and financial support that affected the affordability of women’s care. Such power differences in siblings’ relations oppressed the women regarding care affordability leading to a lack of access to health services.

> *I used to collect vegetables near to my house, but I did not go for asking my brothers to support me. When my parents died, I had a belief that my brothers would not deprive me from properties. Now you see I cannot afford the cost of visiting or staying hospital. I cannot afford even the cost of tests* [pathology] *and medications. I am living with old age allowance from the government, which is not adequate to continue my treatments, but my brothers do not look at my condition*. [Older Woman 8; p.188:1-5]

One staff member spoke about property entitlements between siblings of different genders.

> *Women* [sisters] *do not have the same rights on parents’ property as men* [brothers]. *Some are getting property from father’s house, but rest of the women are denied by their brothers who do not want to give them property rights. It is also a cultural issue where brothers are assumed to be taking care of sisters, but real-life examples are different*. [Clinician 1; p.14:16-17]

#### Neighbourhood challenges

Participants linked growing late life challenges for women to negative community perceptions about ageing and neighbours’ responses to their care needs.

Most participants expressed concern regarding neighbours’ disrespect for elders and feelings of insecurity about ageing. Although staff expressed a common perspective that older women received community respect, some specified disrespect among community people towards older neighbours. They identified a lack of education among neighbours as reason for showing disrespect related to ageism, as one staff member stated in the following extract:

> *It varies family to family. In family with educated members, elderly women used to get respect like other members of the family. As most people are uneducated and very poor in this village, they do not know how to respect and support these women*. [Clinician 10; p.108:20-22]

Participants expressed that worry about ageing developed among older women with multimorbidity because they started considering their future ability to obtain homecare and access treatments. Such worry in old age encouraged women to ignore medical conditions and avoid discussing care needs and use of treatment with neighbours.

> *These women feeling unsecured due to their age … When they become aged, they become unsecured in terms of income and spending. They do not get mental support from the family members as well as their neighbours. In this area, social security becomes insufficient in their age because neighbours are either poor or not supportive to their elderly neighbours*. [Clinician 9; p.93:5-8]
>
> *There is nothing for me. I do not have assets. I feel that I am old who have no rights to live in this society. I am waiting for the call* [death] *from my Allah. I did not get any respect from this community and they do not care about older people. It seems that they will never be at this old age*. [Older Woman 2, p.131: 6-9]

Negative responses from political, economic, and religious elites generated feelings of being an outsider, according to staff and older women, and affected daily activities including healthcare. Political and social elites considered these women unproductive, while religious elites encouraged faith healing, discouraged family members from taking women to formal medical treatment, and deterred women from travelling alone to hospitals where they would see a male doctor.

> *These women do not get enough priority like other population groups in the society. … they cannot go for political movements, so that they are useless for local political leaders. For leaders, elderly women have no value. … political representatives do not support elderly women as they cannot participate in political movements. Political leaders seek young people, who can help them in movements*. [Pharmacist 5; p.84:12-17]
>
> *But most of them are not getting priority in the society. These women cannot contribute in the development of this society as they become aged – this is a common believe in the society*. [Clinician 3; p.33:21-23]
>
> *This is all about religious matter. In Islam, there is restriction for women in going out alone to see a male doctor. We cannot say it as superstition rather this is a religious convention*. (Public Health Assistant 8; p.82:17-19]

Thus, the social alienation of women with multimorbidity because of their age, gender, and economic status hindered them in obtaining support from socio-political and religious elites that could have otherwise have helped them access social and healthcare services and resources.

#### Gender inequality in interactions

Some participants described patriarchal practices in community relationships as leading to women’s poor representation in community activities, restrictions on free movement, and challenges building social relationships. Staff stated that men mainly represented community associations, with older men more highly regarded than older women. Additionally, according to the REW, men’s suppression of women’s relationships continued over a lifetime. Staff described these women as obedient to patriarchal cultural practices that led them to home confinement, which negatively affected their access to local community clinics and hospitals.

> *They have a Pachayet committee (i*.*e*., *village council) where elderly men are involved. Elderly women have no scope to take part in the committee*. [Clinician 2; p.23:12-13]
>
> *I have no relationship with my relatives and neighbours. When my husband died, I lost all relations with them. People used to come to my house regularly. After departing my husband, no-one comes to see me. My husband had a lot of assets, but I do not have anything now. I have no savings or property, so that I am valueless for people. I can remember that my house looked like a marketplace as many people used to gather in my house. We had land and money. Now we have nothing. At present, I have only my Allah and myself*. [Older Woman 14; p.240:6-11]

### Legal connections

The final theme of ‘legal connections’ highlighted a clinician-patient relationship in which older women with multimorbidity often faced clinicians’ ignorance of their rights and missed communication in health facilities.

#### Ignorance of rights

Participants spoke about ineffective health laws and staff abdication of responsibility for meeting the care needs and safety of the older women in hospitals. As aged and multimorbid patients, these women received little or no attention from hospital authorities and staff in relation to upholding their rights to use health services. Although women’s rights were acknowledged, one clinician in the following excerpt discussed how health laws were not practiced. Ignorance of care needs in the hospital system created challenges for women in terms of unequal access.

> *They look like big babies, but they do not receive healthcare like babies. Community clinics or local health centres have special focus on pregnant women and children. What rights can be found for elderly women in Bangladesh? … There is nothing about health rights for elderly women, especially for those who cannot travel to hospital because of their disability or chronic conditions. Health rights are being violated everywhere. Where are the rights for these elderly women in the hospitals? I know there are modified stairs for older patients in developed countries, but here, you cannot even see wheelchairs, welcoming approach or a special care unit for them*. [Clinician 1; p.12:15-19]

Staff rejection of responsibility for women’s care needs was identified as a barrier for adequate and timely use of health services. Most staff expressed a belief that they did not bear any responsibility for older women with multimorbidity, and as such, they denied women’s access to hospital care and/or care at local community clinics.

> *When my son took me to the community clinic* [government subsidised facility], *doctor* [Public Health Assistant who are referred as doctor by most of the women participants] *asked me to meet him in his private clinic in the afternoon. He said that he had no equipment or medication in community clinic for treating me. When I told him about my financial situation, he then asked another lady to talk with me. This lady told that this community clinic was not for older people, so they cannot take the irresponsibility*. [Older Woman 13; p.234:9-12]
>
> *Few days ago, a lady [elderly women with multimorbidity] visited me and asked about her treatment, I told her that you have lots of health problem, thus, you have to go for seeing a doctor in district hospital* [tertiary hospital of the district]. [Clinician 3; p.27:5-6]

Although some staff demonstrated empathy, they still often refused to provide services for these women. The option remained to receive healthcare from doctors and nurses at district level hospitals, but staff’s contribution to poor healthcare experiences discouraged use.

#### Missed communication

Most participants’ views concerning clinicians’ approaches revealed perceptions of older women’s incompetence in communicating with them. This included verbal communication in which staff were unsupportive in response, and a lack of older women’s access to communication technologies such as mobile phones and computers. Thus, family members mediated women’s communication with doctors and nurses, which contributed to further marginalizing them in clinical communication.

> *But sometimes they do not like to hear from me. They said that we know better than you, and you have no need to say anything about your condition. They just give me a prescription without knowing anything about my health problems. These medications do not work for my diseases. They do not like to listen me, and they asked my son to tell them about my diseases. How much I can say to my son about my personal problems [diseases]*. [Older Woman 3; p.139:3-7]
>
> *… doctors and nurses do what they want to do. This is not related with my satisfaction or dissatisfaction. Will they listen to me? Why they listen to a poor and old woman like me? If I cried for the whole day in front of them, they would never listen to me. They will do according to their rules in the hospital*. [Older Woman 8; p.189:4-7]

This lack of communication in combination with staff members’ verbal aggression impacted women’s meaningful use of treatment.

Verbal abuse by staff further discouraged the older women with multimorbidity and their family members from visiting hospitals. Although most clinicians were described as empathetic because of the women’s age, some women reported verbal abuse from medical staff that led them to remain silent during clinical consultation time.

> *They have short temperament. We have to keep patience when they become angry … If you give some money, they will behave very well with you. They will be cool if they get money. Can you understand this? It is all about money. When they see money in patient’s hand, they become greedy. They do not want to lose any chance of taking money from patients*. [Older Woman 12; p.224:5-10]
>
> *… staff behave badly to them. Many poor women came to me and said that the hospitals and clinics staff are shikkhito pagol (i*.*e*., *educated psycho). They are accused to use slang words behind or in front of the women that make them* [women] *shy to show physical symptoms to the doctors and nurses*. [Public Health Assistant 7; p.65:9-10]

Participants’ experiences and perspectives provided insight into several relationship dimensions in older women’s homecare and health service usage to treat multimorbid conditions: a requirement of attention and shared decision making in marital relationships; the focus of family in women’s care needs; ensuring equal rights to parental property to improve care affordability; ageism and disrespect from neighbours; prevalence of gender-based social relationships; medical staff rejection of responsibilities; and passive staff-women relationships in providing care.

## Discussion

The study’s findings fill a gap in the nexus of ageing, care, and dependency by illuminating the meaning of relationships and their dimensions to older women with multimorbidity [Figure 1]. Exploration of interactions in family, neighbourhood and health service spheres is guided by Honneth’s (1996) ‘recognition and misrecognition’ theory that focuses on different facets of relationships, such as personal, family, neighbourhood, and community domains, practice of legal rights, and clinician-patient interactions. Associated events and realities that emerged in this study contribute to the contextualisation of complex relationship dynamics among older women with family members, peers, neighbours, and health staff. Critical social theories and attachment theories suggest that the positive and negative aspects of relationships can influence the health and well-being of older women. The ways that relationship facets shape the homecare and health service use of older women with multimorbidity are discussed in following sections.

Study participants’ insights deepen understanding of constructions of ‘intimacy and care’ in late life. In intimate relationships, spouses, sons/daughters, and in-laws provide a sense of love and non-medical personal care (e.g. meal preparation, bathing, dressing, dish washing, home cleaning, laundry, bed making, emotional support, and management of health service use) to older women with multimorbidity [24, 25, 26]. Personal love and care encourage reliance on family and promote health seeking behaviours. However, poor relationship quality, such as lack of access to family income, male dominance in decision making, mutual disrespect, and intense caregiving for family members, take a toll on an older women’s care needs, especially those with multimorbid conditions. In contrast to research evidence that praises a family care approach, this study reveals the role of family members is highly conditional and not given or fixed in terms of non-medical care [26, 27]. Relationships with spouses [if spouse died - sons, daughters, and in-laws] are of great importance for women’s ability to access family income/savings and make decisions to obtain medical treatment [27, 28]. Literature confirms that older women feel somewhat independent if they can stay in their own homes [29, 30, 31]. However, interviews demonstrate that the home-centredness of older women with multimorbidity is rooted in dependency on family members, where misrecognition often leads a lack of care. Additionally, the structure of intimate relationships may disadvantage women based on gender and socioeconomic status and further the intergenerational transmission of inequality.

This study reveals variation in peer and neighbourhood relationships, based on sociocultural elements, such as education, money and religiosity, in meeting different levels of emotional and financial care needs of older women. Relationships with siblings, friends, neighbors, and community elites have a cumulative influence on health and well-being [31, 32, 33]. Poor peer and neighbourhood relationships challenge women’s ability to obtain health advice, catch-up in sharing problems, accompaniment in travelling, and financial assistance, which are central to the community care [32, 34, 35]. According to this study, as found in other literature, older women ignore and/or delay health service use due to a sense of burden on peers and neighbours. There is consistency in evidence that a lack of education among peers and neighbors prevent them from understanding elder care needs and that strained relationships can impact women’s health outcomes. In a Muslim majority country like Bangladesh, men primarily dominate formal economy and cultural practices, and this imbalance determines women’s community relationships [34, 35, 36]. Older women with multimorbidity either ignore their medical conditions or delay visits to clinics to maintain their dignity in community relationships [37, 38].

Positive relationships between older multimorbid women patients and health staff depend upon recognition of women’s rights, professionalism in care approach, empathy, and shared decision-making. The current landscape of clinical interactions, in combination with limited geriatric services, presents unique challenges and pressures for these patients [35, 39]. Older women with multiple diseases experience complex care needs; however, in the presence of diverse family structures with or without spouses, children and grandchildren, the extent of staff obligations to provide care are less clear [37, 40, 41]. It is vital to address ways to ease service provision and shift burden away from families to health staff through a variety of programs, such as resources for in-home, non-medical care, patient advocates, and the establishment of elder residential villages. These programs can strengthen support structures for older women with multimorbidity to navigate and access hospital care. These women may also experience competing pressures from health staff because of their negative approach and marginalisation in health decision making [42]. Increasing professional responsibility and empathy among staff for patients can improve the quality of clinical consultancy, ensuring older women with multimorbidity receive the full benefit of more positive aspects of service provider-client relationships.

## Limitations

This study has two primary limitations; firstly, the sample of 11 healthcare professionals and 22 older women from one district participated; and secondly, the self-reported accounts of care support by family caregivers, peers, neighbours and health staff is not measured in terms of the diseases and care phases. As a result, the findings may not be generalised, however, these findings may be relevant for project investigating older women in other regions of Bangladesh and developing countries.

## Conclusion

Family, peers, neighbour, and clinical relationships factor into a range of care services and supports for those with multimorbid conditions. Recognition of older multimorbid women patients’ care needs include supporting independence and involvement in the family, community, and hospitals in accessing homecare and health services. These findings demonstrate the importance of family caregivers and staff in espousing a comprehensive care plan and formal recognition of homecare. At the same time, emphasis on relationship dimensions can confirm women’s independence in making health decisions. Peer-education programs for family members and neighbours and a door-to-door health program for older women have potential to change the perception of aging in a developing country like Bangladesh. The care dependency of older women with multimorbidity on different relationships suggests that policy makers need to introduce a holistic care plan, rather than assume that elder care is the family’s responsibility. The study findings highlight the significance of staff training for clinical relationships and increasing knowledge about multimorbid patients’ needs to ensure adequate and timely care for women who must address their chronic health problems while also suffering gender inequality in a social, cultural, religious, and economic context that devalues them in old age.

## Data Availability

All the transcripts [interviews] will be available upon acceptance of the paper

## Acknowledgements

We are thankful to the rural older women and health staff who provided time and shared their experiences about the multimorbid older women patients’ care needs and relationships aspects. We thank Ministry of Health & Family Welfare, Bangladesh; Director of Primary Health Care, Bangladesh; and Sylhet Civil Surgeon Office, Bangladesh for their permission and support in the data collection.

## References

1. Hamiduzzaman M., De Bellis A., Abigail W, Kalaitzidis E. Elderly women in rural Bangladesh: healthcare access and ageing trends. South Asia Research 2018; 38(2): 113–36. doi:10.1177/0262728018767018

2. Hamiduzzaman M. The World is Not Mine: Factors and Issues of Rural Elderly Women’s Access in Modern Healthcare Services in Bangladesh. PhD Thesis, South Australia: Flinders University; 2018.

3. Hossen A, Westhues A. In search of healing between two worlds: the use of traditional and modern health services by older women in rural Bangladesh. Social Work in Health Care 2012; 51(4): 327–44. doi: 10.1080/00981389.2011.638223

4. Ferdous T, Kabir ZN, Wahlin Å, Streatfield K, Cederholm T. The multidimensional background of malnutrition among rural older individuals in Bangladesh – a challenge for the Millennium Development Goal. Public Health Nutrition 2009; 12(12): 2270–278. doi: 10.1017/S1368980009005096

5. Islam MN, Nath DC. A future journey to the elderly support in Bangladesh. Journal of Anthropology 2012; 2012: 1–6. doi: 10.1155/2012/752521

6. Kabir R, Khan H, Kabir M, Rahman T. Population ageing in Bangladesh and its implication on health care. European Scientific Journal 2013; 9(33): 34–47. doi: 10.19044/esj.2013.v9n33p%25p

7. Kabir ZN, Ferdous T, Cederholm T, Khanam MA, Streatfied K, Wahlin Å. Mini nutritional assessment of rural elderly people in Bangladesh: the impact of demographic, socio-economic and health factors. Public Health Nutrition 2006; 9(8): 968–74. doi: 10.1017/PHN2006990

8. Bangladesh Bureau of Statistics. Adjusted Population and Housing Census. Dhaka: Ministry of Planning; 2015.

9. Torres S, Cao X. Improving care for elders who prefer informal spaces to age-separated institutions and health care settings. Innovation in aging. 2019 Jul;3(3):igz019.

10. Hamiduzzaman M, De Bellis A, Kalaitzidis E, Abigail W. Factors impacting on elderly women’s access to healthcare in rural Bangladesh. Indian Journal of Gerontology 2016; 30(2): 235–260.

11. Hossen MA. Older women (User) perspective toward service delivery system of government hospital: a study on some upazila health complex of Bangladesh. Society & Change 2016; 10(1): 18–27.

12. Ferdous T, Cederholm T, Kabir ZN, Hamadani JD, Wahlin Å. Nutritional status and cognitive function in community-living rural Bangladeshi older adults: data from the poverty and health in ageing project. Journal of the American Geriatrics Society 2010; 58(5): 919–24.

13. Hossen A. Bringing Medicine to the Hamlet: Exploring the Experiences of Older Women in Rural Bangladesh who Seek Health Care. PhD thesis. Canada: Wilfrid Laurier University; 2010.

14. Hamiduzzaman M, De Bellis A, Abigail W, Kalaitzidis E. Social determinants of rural elderly women’s healthcare access: a systematic review of qualitative literature. The Indian Journal of Social Work 2018; 39(4): 469–96.

15. Hamiduzzaman M, De Bellis A, Abigail W, Kalaitzidis E. Social determinants for rural elderly women’s access to healthcare: a systematic review of quantitative studies. The Open Public Health Journal 2017; 10(2017): 244–66. doi: 10.2174/1874944501710010244

16. Hossen A, Westhues A. A socially excluded space: restrictions on access to health care for older women in rural Bangladesh. Qualitative Health Research 2010; 20(9): 1192–1201. doi:10.1177/1049732310370695

17. Hossen A, Westhues A. Rural women’s access to health care in Bangladesh: swimming against the tide? Social Work in Public Health 2011; 26(3): 278–93. doi: 10.1080/19371910903126747

18. Hossen A, Westhues A, Maiter S. Coping strategies of older rural Bangladeshi women with health problems. Health Care for Women International 2013; 34(12): 1116–135. doi:10.1080/07399332.2013.817410

19. Thorne S. Interpretive description: qualitative research for applied practice. Routledge, London; 2016.

20. Thorne S, Kirkham SR, MacDonald-Emes J. Focus on qualitative methods. Interpretive description: a non-categorical qualitative alternative for developing nursing knowledge. Research in Nursing & Health 1997; 20(2): 169–77.

21. Braun V, Clarke V. Using thematic analysis in psychology. Qualitative Research in Psychology 2006; 3(2): 77–101. doi: 10.1191/1478088706qp063oa

22. Fairclough N. Critical discourse analysis and critical policy studies. Critical Policy Studies 2013; 7(2): 177–97. doi.org/10.1080/19460171.2013.798239

23. Equator Network. Enhancing the QUAlity and Transparency Of health Research. Consolidated criteria for reporting qualitative research (COREQ): a 32-item checklist for interviews and focus groups (http://www.equator-network.org/reporting-guidelines/coreq/, Accessed 17 May 2018).

24. National Health and Medical Research Council. National statement on ethical conduct in human research, Australian government document; 2015, accessed on 26 May 2018, https://www.google.com.au/url?sa=t&rct=j&q=&esrc=s&source=web&cd=2&cad=rja&uact=8&ved=0ahUKEwjsteHJxZzYAhULerwKHVlmDqoQFgg1MAE&url=https%3A%2F%2Fwww.nhmrc.gov.au%2Fprintpdf%2Fbook%2Fexport%2Fhtml%2F51613&usg=AOvVaw2PVYZwVUA92gLouVQNVkOb.

25. Honneth A. Recognition or redistribution? Theory, Culture & Society 2001; 18(2-3): 43–55. doi:10.1177/02632760122051779

26. Miyawaki CE. A review of ethnicity, culture, and acculturation among Asian caregivers of older adults (2000-2012). SAGE Open 2015; 5(1): 21–58. doi: 10.1177/2158244014566365

27. Sultana AM. Socio-cultural dimensions of women’s discriminations in rural communities. Ozean Journal of Social Sciences 2010; 3(1):31–38.

28. Uddin MT, Islam MN, Alam MJ, Baher GU. Socio-economic status of elderly of Bangladesh: a statistical analysis. Journal of Applied Sciences 2010; 10(23): 3060–67. doi: 10.3923/jas.2010.3060.3067

29. Biswas P, Kabir ZN, Nilsson J, Zaman S. Dynamics of health care seeking behaviour of elderly people in rural Bangladesh. International Journal of Ageing and Later Life 2006; 1(1): 69–89. doi: 10.3384/ijal.1652-8670.061169

30. Abdulraheem IS. Health needs assessment and determinants of health-seeking behaviour among elderly Nigerians: a house-hold survey. Annals of African Medicine 2007; 6(2): 58–63. doi: 10.4103/1596-3519.55715

31. Ahern A, Hine J. Accessibility of health services for aged people in rural Ireland, International Journal of Sustainable Transportation 2015; 9(5): 389–95. doi: 10.1080/15568318.2013.800926

32. Alam N, Barkat-e-Khuda Demographic events and economic conditions of rural households in Bangladesh. Asian Population Studies 2014; 10(3): 304–18. doi: 10.1080/17441730.2014.890162

33. Lussier DN, Fish MS. Men, Muslims, and attitudes toward gender inequality. Politics and Religion 2016; 9(1): 29–60.

34. Hegland ME. Gender and religion in the Middle East and South Asia: Women’s voices rising. In A social history of women and gender in the modern Middle East (pp. 177-212). Routledge; 2018.

35. Allen L, Williams J, Townsend N, Mikkelsen B, Roberts N, Foster C, Wickramasinghe K. Socioeconomic status and non-communicable disease behavioural risk factors in low-income and lower-middle-income countries: a systematic review. The Lancet Global Health 2017; 5(3): 277–289. doi: 10.1016/S2214-109X(17)30058-X

36. Ameh S, Gomez-Olive FX, Kahn K, Tollman SM, Klipstein-Grobusch K. Predictors of health care use by adults 50 years and over in a rural South African setting. Global Health Action 2014; 7(1): 1–11. doi: 10.3402/gha.v7.24771

37. Amin I. Perceptions of successful aging among older adults in Bangladesh: an exploratory study. Journal of Cross-cultural Gerontology 2017; 32(2): 191–207. doi: 10.1007/s10823-017-9319-3

38. Bergeron CD, Hilfinger-Messias DK, Friedman DB, Spencer SM, Miller SC. Involvement of family members and professionals in older women’s post-fall decision making. Health Communication 2018; 33(3): 246–53. doi: 10.1080/10410236.2016.1255844

39. Begum S, Wesumperuma D. Overview of the Old Age Allowance Programme in Bangladesh. In Social Protection for Older Persons: Social Pensions in Asia. Eds by S. W. Handayani & B. Babajanian. Philippines: Asian Development Bank; 2012.

40. Huang H, Sharifian F, Feldman S, Yang H, Radermacher H, Browning C. Cross-cultural conceptualizations of ageing in Australia. Cognitive Linguistic Studies 2018; 5(2): 261–81.

41. Rahman MM. Health status and health needs among the aged population in Chapai Nawabganj district of Bangladesh. Indian Journal of Gerontology 2009; 23(1): 32–41.

42. Hamiduzzaman M. Self-reported seasonal symptoms and diseases and primary healthcare utilization among rural elderly women in Sylhet District, Bangladesh. Journal of UoEH 2020; 42(2): 175–185.

